# Comparison of Transcatheter Edge-to-Edge Mitral Valve Repair for Primary Mitral Regurgitation Outcomes to Hospital Volumes of Surgical Mitral Valve Repair

**DOI:** 10.1101/2023.06.19.23291628

**Authors:** Paul A. Grayburn, Michael J. Mack, Pratik Manandhar, Andrzej S. Kosinski, Anna Sannino, Robert L. Smith, Molly Szerlip, Sreekanth Vemulapalli

## Abstract

**Background:** Transcatheter edge-to-edge mitral valve (MV) repair (TEER) is an effective treatment for patients with primary mitral regurgitation (MR) at prohibitive risk for surgical MV repair (MVr). High volume MVr centers and high volume TEER centers have better outcomes than low volume centers, respectively. However little is known about whether MVr volume, and specifically complex MVr volume, predicts TEER outcomes. We hypothesized that high volume MV surgical centers would have superior risk-adjusted outcomes for TEER than tlow volume centers.

**Methods:** We combined data from the ACC/STS TVT registry and the STS adult cardiac surgery database. Complex MVr, defined as leaflet resection or artificial chords with or without annuloplasty was evaluated as a continuous variable and as pre-defined categories (<25, 25-49 and ≥ 50 MV repairs/year). A generalized linear mixed model was used to evaluate risk-adjusted in-hospital/30-day mortality, 30 day HF readmission and TEER success (MR ≤ 2+ and gradient < 5 mmHg).

**Results:** The study comprised 41,834 patients from 500 sites. TEER mortality at 30-days was 3.5% with no significant difference across MVr volume on unadjusted (p=0.141) or adjusted (p=0.071) analysis of volume as a continuous variable. One-year mortality was 15.0% and was lower for higher MVr volume centers when adjusted for clinical and demographic variables (p=0.027). HF readmission at one year was 9.4% and was statistically significantly lower in high volume centers on both unadjusted (p=0.017) or adjusted (p-0.015) analysis. TEER success was 54.6% and was not statistically significantly different across MV surgical site volumes (p=0.4271).

**Conclusions:** TEER can be safely performed in centers with low volumes of complex MV repair. However, one-year mortality and HF readmission are superior at centers with higher MVr volume.

---

Transcatheter edge-to-edge mitral valve (MV) repair (TEER) with MitraClip^TM^ (Abbott Vascular Structural Heart, Santa Clara, California) has emerged as an effective treatment for eligible patients with mitral regurgitation (MR). MitraClip^TM^ has been commercially available in the United States since 2013 for patients with primary degenerative MR, appropriate anatomy, and prohibitive risk for MV surgery (1,2). It was approved in 2018 for secondary MR in patients with persistent severe MR despite guideline-directed medical therapy with left ventricular ejection fraction between 20-50% based on results from the COAPT trial (3). TEER is a complex procedure wherein multiple factors including operator and echocardiographer experience, patient selection, and MV anatomy may affect procedural success. Prior observational studies from the Society of Thoracic Surgeons/American College of Cardiology Transcatheter Valve Therapy Registry (STS/ACC TVT registry) have demonstrated that procedural outcomes of TEER improve with increasing institutional as well as operator experience (4,5). Badhwar et al (6) examined data from the STS Adult Cardiac Surgery Database to assess whether procedural volume influences the outcomes of MV surgery. They reported that volume-outcome relationship exists at both the hospital and surgeon levels for operative mortality, 1-year mortality, 30-day composite of mortality/morbidity, and rate of successful repair. High volume hospitals and surgeons have demonstrated superior outcomes compared to the low volume hospitals and surgeons. The annual hospital volume appeared to flatten out at around 75 cases at the hospital level and 35 cases at the surgeon level (6).

Hirji et al (7) demonstrated that lower hospital surgical aortic valve replacement volume was correlated with transcatheter aortic valve replacement (TAVR) outcomes, with higher 30-day and 1-year mortality observed at low-volume centers. As a more complex procedure than TAVR, TEER relies heavily on the collaboration among multidisciplinary specialists for patient selection and procedural performance. Current Medicare coverage policy requires that hospitals perform ≥25 annual MV surgeries before starting a TEER program. However, Barker et al. (8), in a claims-based single year data analysis, concluded that for TEER patients, mortality and cardiac hospitalization rates did not vary significantly by institutional volumes for either MV surgery or TEER. This lack of a difference in outcomes based on TEER volume could be explained by the low incidence of in-hospital mortality and small number of procedures included in the study. We hypothesized that hospital MV surgical volumes are associated with TEER outcomes. We anticipated that high volume MV surgical centers would have higher STS predicted risk of mortality for MV repair than lower volume centers but with superior risk-adjusted outcomes. To evaluate this hypothesis, we combined data from both the ACC/STS TVT registry and the STS adult cardiac surgery database.

## METHODS

### Patient Population and Study Design

We included patients who underwent TEER with MitraClip^TM^ from 1/1/2014 to 12/31/2021 in ACC/STS TVT Registry. The STS/ACC TVT Registry contains nearly all commercial transcatheter mitral procedures done in the United States, as required by the Centers for Medicare and Medicaid Services coverage decision. The registry undergoes data quality and completeness checks at both the American College of Cardiology as well as the Duke Clinical Research Institute. To further ensure data quality and completeness, the registry undergoes a random 10% yearly audit by a 3^rd^ party. We restricted the analysis to patients with primary MR (prolapse/flail leaflets) for three reasons. First, primary and secondary MR are different disease entities with differing natural history and treatment (9). Second, the standard treatment for primary MR due to prolapse or flail leaflets is surgical repair (when feasible) or TEER when surgical risk is prohibitive. Third, primary MR (Carpentier Type II) is more often correctly classified by cardiologists and cardiac surgeons than secondary MR which is sometimes misclassified as prolapse due to anterior leaflet override or misjudged as secondary (Carpentier type IIIB) when it is in fact primary MV leaflet restriction (Carpentier Type IIIA) (10). Otherwise, there were no exclusion criteria.

### Institutional Complex MVr volume

Hospital NPI number of Institutions performing TEER in the STS/ACC TVT Registry were identified. These NPI numbers were used to identify the same institutions within the STS Adult Cardiac Surgery Database. Complex surgical valve cases were defined in the Adult Cardiac Surgery Database as as leaflet resection or artificial chords with or without annuloplasty or valve replacement, excluding annuloplasty only. Annualized surgical MVr volume was calculated as (365 x Total number of surgical MVr) / (Last surgical MVr date – First surgical MVr date + 1)

### Endpoint Definitions

The primary endpoint was in-hospital or 30-day and 1-year mortality after TEER. The secondary endpoints were 30-day and 1 year heart failure (HF) readmissions and TEER success. Heart Failure hospitalization was defined as a hospital or ER stay of >/=24 hours with treatment for heart failure. TEER success was defined as residual MR grade ≤2+ with a mean mitral gradient < 5 mmHg; optimal device success was defined as residual MR grade ≤1+ with a mean mitral gradient < 5 mmHg.

### Statistical Analysis

A pre-specified analysis plan was derived to assess the relationship between TEER outcomes and MV surgical volumes. Hospital MVr volume was evaluated using two different methods: as a continuous variable with restricted cubic splines for nonlinearity and additionally as pre-defined categories (<25, 25-49 and ≥ 50 MV surgeries/year). Generalized linear mixed models were used to evaluate risk-adjusted in-hospital/30-day mortality, 30 day HF readmission and MV repair success rates. Variables used for adjustment are listed in Supplemental Table 1. The model for TEER success additionally adjusts for site region, teaching status, number of beds and community. The 2-level hierarchical structure of the data with patients nested within sites and random residuals was used to account for clustering within sites. One-year all-cause death was assessed using Cox proportional hazards model and one-year HF Readmission was assessed using Fine and Gray’s sub-distribution hazards model. To account for bias due to missing data on one-year outcomes, inverse proportional weights (IPW) approach was used. Under the IPW approach, a logistic regression was first used to calculate the probability of having non-missing data on one-year outcome and subsequent survival model was then weighted on the inverse propensity of having non-missing one-year outcomes. Continuous variables were presented as as median (interquartile range), and categorical variables as the percentage (frequency), unless otherwise specified. Categorical variables were compared using the chi-square or Fisher exact test, as appropriate. Continuous variables were compared using Kruskal-Wallis test. A 2-sided 5% significance level was used throughout. Statistical calculations were performed using SAS 9.4 (Cary, NC, USA). All analyses were done at the Duke Clinical Research Institute. The STS/TVT Registry has approval through the Advarra IRB and this analysis was granted a waiver by the Duke University Institutional Review Board.

## Results

Fig. 1 shows the flow chart for patient inclusion. Of 54,422 patients undergoing TEER at 514 sites during the study period, 41,834 patients from 500 sites were included. Patients with non-primary MR (n=12,051) and patients with no matching records in the STS database (n=537) were excluded. Table 1 shows baseline characteristics for the overall cohort and each of the MV surgical groups. Although many variables were statistically significant between groups of high, intermediate and low surgical volume centers due to the high numbers of subjects, most were not clinically significant. Examples include age (median 80, 81, 81, p<0.001) and serum creatinine (median 1.2, 1.2, 1.2, p<0.001). Of potential relevance, two or more prior cardiac surgeries and prior aortic valve intervention were slightly more common in high volume centers. STS predicted risk of mortality for MV repair was higher in high volume centers (4.7%, 5.1%, 5.5%, p<0.0001). Moderate or severe tricuspid regurgitation was common and slightly higher in high MVr volume centers (median 46.4, 47.0, 49.0, p<0.001). Failure to document location of prolapse (25.3% overall) or flail leaflet (37.1%) was common. Of the 500 sites, 332 (66.4%) were low, 102 (20.4%) intermediate and 66 (13.2%) high volume surgical centers (p<0.001). There were regional differences across surgical volume centers and teaching hospitals and number of hospital beds were significantly higher in high volume centers.

**Fig 1.**
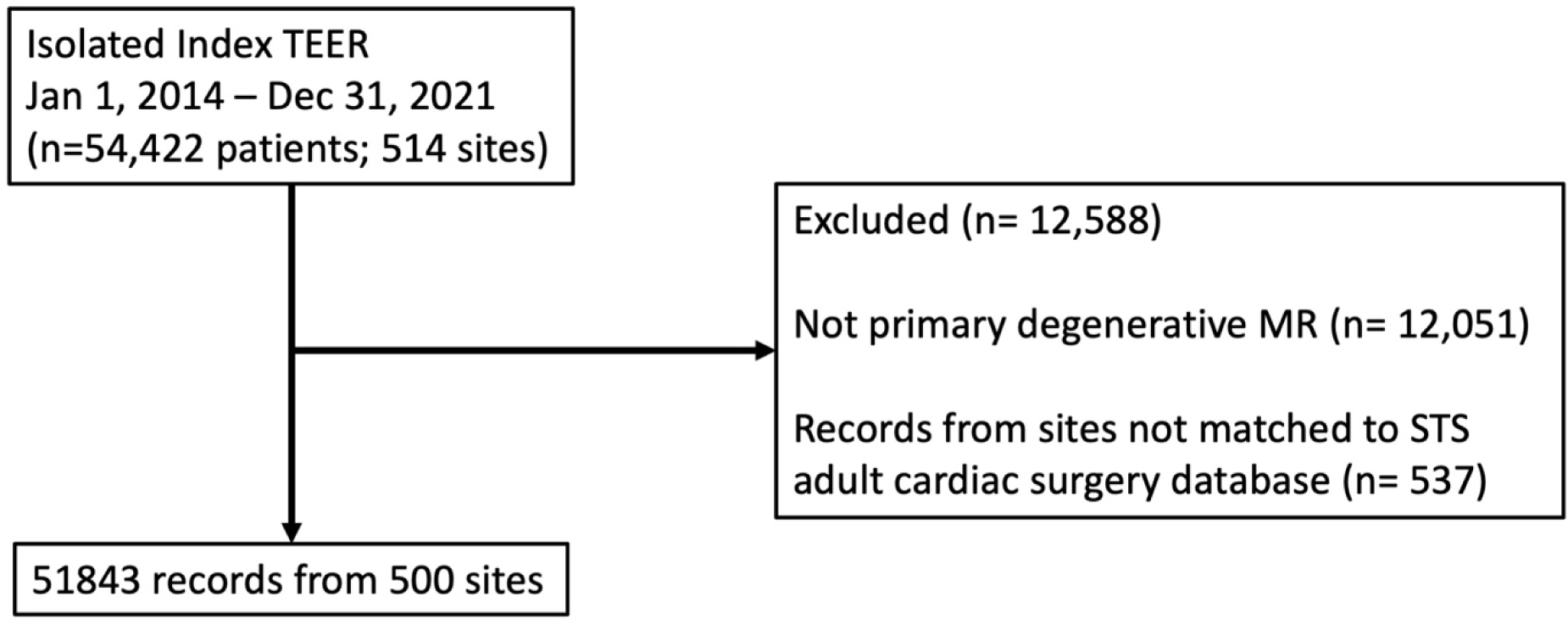
Flow diagram.

**Table 1.** Patient baseline demographics by annualized surgical MV volume.

| Variable | Level | Overall |  | Surgical MV<br>volume < 25<br>[Sites = 332] |  | Surgical MV<br>volume 25 - 49<br>[Sites = 102] |  | Surgical MV<br>volume 50+<br>[Sites = 66] |  | P-<br>value+ |
| --- | --- | --- | --- | --- | --- | --- | --- | --- | --- | --- |
|  |  | (N=41834) |  | (N=19778) |  | (N=11318) |  | (N=10738) |  |  |
| **<u>Demographics</u>** |  |  |  |  |  |  |  |  |  |  |
| Age++ | Median | 41834 | 80.0 | 19778 | 80.0 | 11318 | 81.0 | 10738 | 81.0 | <.0001 |
|  | 25th |  | 73.0 |  | 73.0 |  | 74.0 |  | 74.0 |  |
|  | 75th |  | 85.0 |  | 85.0 |  | 86.0 |  | 86.0 |  |
|  | Missing(%) |  | 0.0 |  | 0.0 |  | 0.0 |  | 0.0 |  |
| BSA++ | Median | 41757 | 1.8 | 19733 | 1.8 | 11298 | 1.8 | 10726 | 1.8 | <.0001 |
|  | 25th |  | 1.7 |  | 1.7 |  | 1.7 |  | 1.6 |  |
|  | 75th |  | 2.0 |  | 2.0 |  | 2.0 |  | 2.0 |  |
|  | Missing(%) |  | 0.2 |  | 0.2 |  | 0.2 |  | 0.1 |  |
| Gender | Female | 19474 | 46.6 | 9414 | 47.6 | 5306 | 46.9 | 4754 | 44.3 | <.0001 |
|  | Male | 22359 | 53.4 | 10363 | 52.4 | 6012 | 53.1 | 5984 | 55.7 |  |
|  | Missing | 1 | 0.0 | 1 | 0.0 | 0 | 0.0 | 0 | 0.0 |  |
| Race-White | Yes | 36931 | 88.3 | 17481 | 88.4 | 9928 | 87.7 | 9522 | 88.7 | 0.0711 |
|  | No | 4903 | 11.7 | 2297 | 11.6 | 1390 | 12.3 | 1216 | 11.3 |  |
| Hispanic or Latino<br>Ethnicity | Yes | 1998 | 4.8 | 1184 | 6.0 | 424 | 3.7 | 390 | 3.6 | <.0001 |
|  | No | 39282 | 93.9 | 18386 | 93.0 | 10757 | 95.0 | 10139 | 94.4 |  |
|  | Missing | 554 | 1.3 | 208 | 1.1 | 137 | 1.2 | 209 | 1.9 |  |
| **<u>History and Risk</u>**<br>**<u>Factors</u>** |  |  |  |  |  |  |  |  |  |  |
| NYHA IV | Yes | 7963 | 19.0 | 3685 | 18.6 | 2182 | 19.3 | 2096 | 19.5 | 0.0990 |
|  | No | 33467 | 80.0 | 15926 | 80.5 | 9012 | 79.6 | 8529 | 79.4 |  |
|  | Missing | 404 | 1.0 | 167 | 0.8 | 124 | 1.1 | 113 | 1.1 |  |
| Pre-procedure<br>Creatinine++ | Median | 41575 | 1.2 | 19667 | 1.2 | 11264 | 1.2 | 10644 | 1.2 | 0.0062 |
|  | 25th |  | 0.9 |  | 0.9 |  | 0.9 |  | 0.9 |  |
|  | 75th |  | 1.5 |  | 1.5 |  | 1.5 |  | 1.6 |  |
|  | Missing(%) |  | 0.6 |  | 0.6 |  | 0.5 |  | 0.9 |  |
| Prior PAD | Yes | 6889 | 16.5 | 3209 | 16.2 | 1933 | 17.1 | 1747 | 16.3 | 0.1156 |
|  | No | 34887 | 83.4 | 16542 | 83.6 | 9365 | 82.7 | 8980 | 83.6 |  |
|  | Missing | 58 | 0.1 | 27 | 0.1 | 20 | 0.2 | 11 | 0.1 |  |
| Hypertension | Yes | 36011 | 86.1 | 17059 | 86.3 | 9754 | 86.2 | 9198 | 85.7 | 0.3492 |
|  | No | 5799 | 13.9 | 2710 | 13.7 | 1556 | 13.7 | 1533 | 14.3 |  |
|  | Missing | 24 | 0.1 | 9 | 0.0 | 8 | 0.1 | 7 | 0.1 |  |
| GFR++ | Median | 41572 | 53.3 | 19664 | 53.7 | 11264 | 53.1 | 10644 | 53.0 | 0.0211 |
|  | 25th |  | 39.0 |  | 39.2 |  | 39.2 |  | 38.5 |  |
|  | 75th |  | 69.5 |  | 69.7 |  | 69.7 |  | 68.8 |  |
|  | Missing(%) |  | 0.6 |  | 0.6 |  | 0.5 |  | 0.9 |  |
| Currently on Dialysis | Yes | 1546 | 3.7 | 761 | 3.8 | 394 | 3.5 | 391 | 3.6 | 0.2406 |
|  | No | 40245 | 96.2 | 18995 | 96.0 | 10914 | 96.4 | 10336 | 96.3 |  |
|  | Missing | 43 | 0.1 | 22 | 0.1 | 10 | 0.1 | 11 | 0.1 |  |
| LVEF++ | Median | 41045 | 55.0 | 19441 | 55.0 | 11132 | 55.0 | 10472 | 57.0 | <.0001 |
|  | 25th |  | 40.0 |  | 40.0 |  | 42.0 |  | 45.0 |  |
|  | 75th |  | 60.0 |  | 60.0 |  | 60.0 |  | 60.0 |  |

| Variable | Level | Overall<br>(N=41834) |  | Surgical MV<br>volume < 25<br>[Sites = 332]<br>(N=19778) |  | Surgical MV<br>volume 25 - 49<br>[Sites = 102]<br>(N=11318) |  | Surgical MV<br>volume 50+<br>[Sites = 66]<br>(N=10738) |  | P-<br>value+ |
| --- | --- | --- | --- | --- | --- | --- | --- | --- | --- | --- |
|  | Missing(%) |  | 1.9 |  | 1.7 |  | 1.6 |  | 2.5 |  |
| Hemoglobin++ | Median | 41530 | 12.2 | 19655 | 12.3 | 11245 | 12.2 | 10630 | 12.0 | <.0001 |
|  | 25th |  | 10.8 |  | 10.9 |  | 10.8 |  | 10.6 |  |
|  | 75th |  | 13.5 |  | 13.5 |  | 13.4 |  | 13.3 |  |
|  | Missing(%) |  | 0.7 |  | 0.6 |  | 0.6 |  | 1.0 |  |
| Left Main Stenosis >= 50% | Yes | 2653 | 6.3 | 1267 | 6.4 | 720 | 6.4 | 666 | 6.2 | 0.9130 |
|  | No | 37269 | 89.1 | 17828 | 90.1 | 9986 | 88.2 | 9455 | 88.1 |  |
|  | Missing | 1912 | 4.6 | 683 | 3.5 | 612 | 5.4 | 617 | 5.7 |  |
| Prior MI | Yes | 9995 | 23.9 | 4671 | 23.6 | 2845 | 25.1 | 2479 | 23.1 | 0.0008 |
|  | No | 31793 | 76.0 | 15085 | 76.3 | 8461 | 74.8 | 8247 | 76.8 |  |
|  | Missing | 46 | 0.1 | 22 | 0.1 | 12 | 0.1 | 12 | 0.1 |  |
| Endocarditis | Yes | 587 | 1.4 | 241 | 1.2 | 168 | 1.5 | 178 | 1.7 | 0.0054 |
|  | No | 41193 | 98.5 | 19515 | 98.7 | 11129 | 98.3 | 10549 | 98.2 |  |
|  | Missing | 54 | 0.1 | 22 | 0.1 | 21 | 0.2 | 11 | 0.1 |  |
| Prior TIA/Stroke | Yes | 6673 | 16.0 | 3055 | 15.4 | 1903 | 16.8 | 1715 | 16.0 | 0.0062 |
|  | No | 35091 | 83.9 | 16692 | 84.4 | 9392 | 83.0 | 9007 | 83.9 |  |
|  | Missing | 70 | 0.2 | 31 | 0.2 | 23 | 0.2 | 16 | 0.1 |  |
| Carotid Stenosis | Yes | 4310 | 10.3 | 2051 | 10.4 | 1222 | 10.8 | 1037 | 9.7 | <.0001 |
|  | No | 23066 | 55.1 | 11320 | 57.2 | 5695 | 50.3 | 6051 | 56.4 |  |
|  | Missing | 14458 | 34.6 | 6407 | 32.4 | 4401 | 38.9 | 3650 | 34.0 |  |
| Peripheral Arterial Disease | Yes | 6889 | 16.5 | 3209 | 16.2 | 1933 | 17.1 | 1747 | 16.3 | 0.1156 |
|  | No | 34887 | 83.4 | 16542 | 83.6 | 9365 | 82.7 | 8980 | 83.6 |  |
|  | Missing | 58 | 0.1 | 27 | 0.1 | 20 | 0.2 | 11 | 0.1 |  |
| Current Smoker | Yes | 2731 | 6.5 | 1407 | 7.1 | 745 | 6.6 | 579 | 5.4 | <.0001 |
|  | No | 38963 | 93.1 | 18301 | 92.5 | 10531 | 93.0 | 10131 | 94.3 |  |
|  | Missing | 140 | 0.3 | 70 | 0.4 | 42 | 0.4 | 28 | 0.3 |  |
| Diabetes | Yes | 11060 | 26.4 | 5343 | 27.0 | 3106 | 27.4 | 2611 | 24.3 | <.0001 |
|  | No | 30711 | 73.4 | 14404 | 72.8 | 8195 | 72.4 | 8112 | 75.5 |  |
|  | Missing | 63 | 0.2 | 31 | 0.2 | 17 | 0.2 | 15 | 0.1 |  |
| Atrial Fibrillation/Flutter | Yes | 26202 | 62.6 | 12107 | 61.2 | 7272 | 64.3 | 6823 | 63.5 | <.0001 |
|  | No | 15578 | 37.2 | 7648 | 38.7 | 4030 | 35.6 | 3900 | 36.3 |  |
|  | Missing | 54 | 0.1 | 23 | 0.1 | 16 | 0.1 | 15 | 0.1 |  |
| Chronic Lung Disease | Severe | 3790 | 9.1 | 1739 | 8.8 | 1189 | 10.5 | 862 | 8.0 | <.0001 |
|  | None/Mild/Moderate | 37024 | 88.5 | 17514 | 88.6 | 9901 | 87.5 | 9609 | 89.5 |  |
|  | Missing | 1020 | 2.4 | 525 | 2.7 | 228 | 2.0 | 267 | 2.5 |  |
| Home Oxygen | Yes | 4740 | 11.3 | 2216 | 11.2 | 1356 | 12.0 | 1168 | 10.9 | 0.0259 |
|  | No | 37058 | 88.6 | 17548 | 88.7 | 9950 | 87.9 | 9560 | 89.0 |  |
|  | Missing | 36 | 0.1 | 14 | 0.1 | 12 | 0.1 | 10 | 0.1 |  |
| Hostile Chest | Yes | 2938 | 7.0 | 1297 | 6.6 | 870 | 7.7 | 771 | 7.2 | 0.0007 |
|  | No | 38864 | 92.9 | 18469 | 93.4 | 10439 | 92.2 | 9956 | 92.7 |  |
|  | Missing | 32 | 0.1 | 12 | 0.1 | 9 | 0.1 | 11 | 0.1 |  |
| Porcelain Aorta | Yes | 413 | 1.0 | 168 | 0.8 | 117 | 1.0 | 128 | 1.2 | 0.0129 |
|  | No | 41349 | 98.8 | 19580 | 99.0 | 11174 | 98.7 | 10595 | 98.7 |  |
|  | Missing | 72 | 0.2 | 30 | 0.2 | 27 | 0.2 | 15 | 0.1 |  |
| Pacemaker | Yes | 7497 | 17.9 | 3622 | 18.3 | 2038 | 18.0 | 1837 | 17.1 | 0.0273 |
|  | No | 34259 | 81.9 | 16116 | 81.5 | 9252 | 81.7 | 8891 | 82.8 |  |
|  | Missing | 78 | 0.2 | 40 | 0.2 | 28 | 0.2 | 10 | 0.1 |  |
| Previous ICD | Yes | 5476 | 13.1 | 2804 | 14.2 | 1451 | 12.8 | 1221 | 11.4 | <.0001 |
|  | No | 36313 | 86.8 | 16952 | 85.7 | 9854 | 87.1 | 9507 | 88.5 |  |
|  | Missing | 45 | 0.1 | 22 | 0.1 | 13 | 0.1 | 10 | 0.1 |  |
| Prior PCI | Yes | 12419 | 29.7 | 6097 | 30.8 | 3270 | 28.9 | 3052 | 28.4 | <.0001 |
|  | No | 29361 | 70.2 | 13660 | 69.1 | 8032 | 71.0 | 7669 | 71.4 |  |
|  | Missing | 54 | 0.1 | 21 | 0.1 | 16 | 0.1 | 17 | 0.2 |  |
| Prior CABG | Yes | 9560 | 22.9 | 4414 | 22.3 | 2625 | 23.2 | 2521 | 23.5 | 0.0438 |
|  | No | 32198 | 77.0 | 15325 | 77.5 | 8674 | 76.6 | 8199 | 76.4 |  |
|  | Missing | 76 | 0.2 | 39 | 0.2 | 19 | 0.2 | 18 | 0.2 |  |
| Number of previous<br>cardiac surgeries | 2+ | 1675 | 4.0 | 681 | 3.4 | 466 | 4.1 | 528 | 4.9 | <.0001 |
|  | 1 | 10525 | 25.2 | 4863 | 24.6 | 2866 | 25.3 | 2796 | 26.0 |  |
|  | 0 | 29205 | 69.8 | 14051 | 71.0 | 7845 | 69.3 | 7309 | 68.1 |  |
|  | Missing | 429 | 1.0 | 183 | 0.9 | 141 | 1.2 | 105 | 1.0 |  |
| Prior Aortic Valve<br>Procedure | Yes | 4129 | 9.9 | 1752 | 8.9 | 1192 | 10.5 | 1185 | 11.0 | <.0001 |
|  | No | 37633 | 90.0 | 17996 | 91.0 | 10103 | 89.3 | 9534 | 88.8 |  |
|  | Missing | 72 | 0.2 | 30 | 0.2 | 23 | 0.2 | 19 | 0.2 |  |
| Pre-Procedure Mod/Sev<br>Aortic Insufficiency | Moderate/Severe | 3397 | 8.1 | 1658 | 8.4 | 925 | 8.2 | 814 | 7.6 | 0.0464 |
|  | None/Trace/Mild | 38081 | 91.0 | 17948 | 90.7 | 10296 | 91.0 | 9837 | 91.6 |  |
|  | Missing | 356 | 0.9 | 172 | 0.9 | 97 | 0.9 | 87 | 0.8 |  |
| Pre-Procedure Tricuspid<br>Insufficiency | Moderate/Severe | 19744 | 47.2 | 9172 | 46.4 | 5314 | 47.0 | 5258 | 49.0 | <.0001 |
|  | None/Trace/Mild | 21756 | 52.0 | 10463 | 52.9 | 5912 | 52.2 | 5381 | 50.1 |  |
|  | Missing | 334 | 0.8 | 143 | 0.7 | 92 | 0.8 | 99 | 0.9 |  |
| Acuity | Emergency/Salvage/C<br>ardiacArrest | 377 | 0.9 | 148 | 0.7 | 110 | 1.0 | 119 | 1.1 | <.0001 |
|  | Shock/Intrope/Device<br>Assist | 2514 | 6.0 | 1030 | 5.2 | 650 | 5.7 | 834 | 7.8 |  |
|  | Urgent | 3572 | 8.5 | 1614 | 8.2 | 925 | 8.2 | 1033 | 9.6 |  |
|  | Elective | 35371 | 84.6 | 16986 | 85.9 | 9633 | 85.1 | 8752 | 81.5 |  |
| STS risk score - MV<br>Repair++ | Median | 41828 | 5.0 | 19773 | 4.7 | 11317 | 5.1 | 10738 | 5.5 | <.0001 |
|  | 25th |  | 3.0 |  | 2.7 |  | 3.1 |  | 3.2 |  |
|  | 75th |  | 8.4 |  | 7.9 |  | 8.6 |  | 9.0 |  |
|  | Missing(%) |  | 0.0 |  | 0.0 |  | 0.0 |  | 0.0 |  |
| Degenerative MR-Leaflet Prolapse | Not Documented | 10589 | 25.3 | 5192 | 26.3 | 3013 | 26.6 | 2384 | 22.2 | <.0001 |
|  | Bileaflet | 4734 | 11.3 | 2009 | 10.2 | 1333 | 11.8 | 1392 | 13.0 |  |
|  | Posterior | 11432 | 27.3 | 5012 | 25.3 | 3173 | 28.0 | 3247 | 30.2 |  |
|  | Anterior | 6225 | 14.9 | 2939 | 14.9 | 1708 | 15.1 | 1578 | 14.7 |  |
|  | None | 8053 | 19.2 | 4159 | 21.0 | 1974 | 17.4 | 1920 | 17.9 |  |
|  | Missing | 801 | 1.9 | 467 | 2.4 | 117 | 1.0 | 217 | 2.0 |  |
| Degenerative MR-Leaflet Flail | Not Documented | 15535 | 37.1 | 7649 | 38.7 | 4556 | 40.3 | 3330 | 31.0 | <.0001 |
|  | Bileaflet | 566 | 1.4 | 250 | 1.3 | 133 | 1.2 | 183 | 1.7 |  |
|  | Posterior | 9640 | 23.0 | 3806 | 19.2 | 2526 | 22.3 | 3308 | 30.8 |  |
|  | Anterior | 2873 | 6.9 | 1161 | 5.9 | 726 | 6.4 | 986 | 9.2 |  |

| Variable | Level | Overall |  | Surgical MV<br>volume < 25<br>[Sites = 332]<br>(N=19778) |  | Surgical MV<br>volume 25 - 49<br>[Sites = 102]<br>(N=11318) |  | Surgical MV<br>volume 50+<br>[Sites = 66]<br>(N=10738) |  | P-<br>value+ |
| --- | --- | --- | --- | --- | --- | --- | --- | --- | --- | --- |
|  |  | (N=41834) |  |  |  |  |  |  |  |  |
|  | None | 12222 | 29.2 | 6321 | 32.0 | 3223 | 28.5 | 2678 | 24.9 |  |
|  | Missing | 998 | 2.4 | 591 | 3.0 | 154 | 1.4 | 253 | 2.4 |  |
| **<u>Site Characteristics</u>** |  |  |  |  |  |  |  |  |  |  |
| Region | South | 193 | 38.6 | 134 | 40.4 | 36 | 35.3 | 23 | 34.8 | <.0001 |
|  | Midwest | 118 | 23.6 | 83 | 25.0 | 21 | 20.6 | 14 | 21.2 |  |
|  | Northeast | 78 | 15.6 | 34 | 10.2 | 22 | 21.6 | 22 | 33.3 |  |
|  | West | 111 | 22.2 | 81 | 24.4 | 23 | 22.5 | 7 | 10.6 |  |
| Teaching Hospital | Yes | 312 | 62.4 | 189 | 56.9 | 71 | 69.6 | 52 | 78.8 | 0.0009 |
|  | No | 188 | 37.6 | 143 | 43.1 | 31 | 30.4 | 14 | 21.2 |  |
| Community Description | Rural | 46 | 9.2 | 38 | 11.4 | 6 | 5.9 | 2 | 3.0 | 0.0028 |
|  | Suburban | 140 | 28.0 | 104 | 31.3 | 25 | 24.5 | 11 | 16.7 |  |
|  | Urban | 314 | 62.8 | 190 | 57.2 | 71 | 69.6 | 53 | 80.3 |  |
| Hospital Beds++ | Median | 500 | 478.0 | 332 | 410.5 | 102 | 547.0 | 66 | 710.0 | <.0001 |
|  | 25th |  | 337.5 |  | 311.5 |  | 440.0 |  | 489.0 |  |
|  | 75th |  | 657.5 |  | 572.0 |  | 700.0 |  | 950.0 |  |
|  | Missing(%) |  | 0.0 |  | 0.0 |  | 0.0 |  | 0.0 |  |

### Mortality (Primary Endpoint

Figure 2A shows restricted cubic spline analysis for in-hospital or 30-day mortality with no statistically significant difference on unadjusted (p=0.141) or adjusted (p=0.071) analysis. Table 2 shows outcomes overall and across the different tiers of surgical volumes. In-hospital or 30-day mortality was 3.5% overall and was not statistically significantly different across MV surgical volumes at 3.4%, 3.4% and 3.9% in MV surgical sites with <25, 25-49 and ≥ 50 annualized cases (p=0.0882). When adjusted for clinical and demographic variables, the relationship between annualized surgical MVr volume and in-hospital and 30-day mortality remained non-significant (p=0.552, Supplemental Table 2A).

**Figure 2a.**
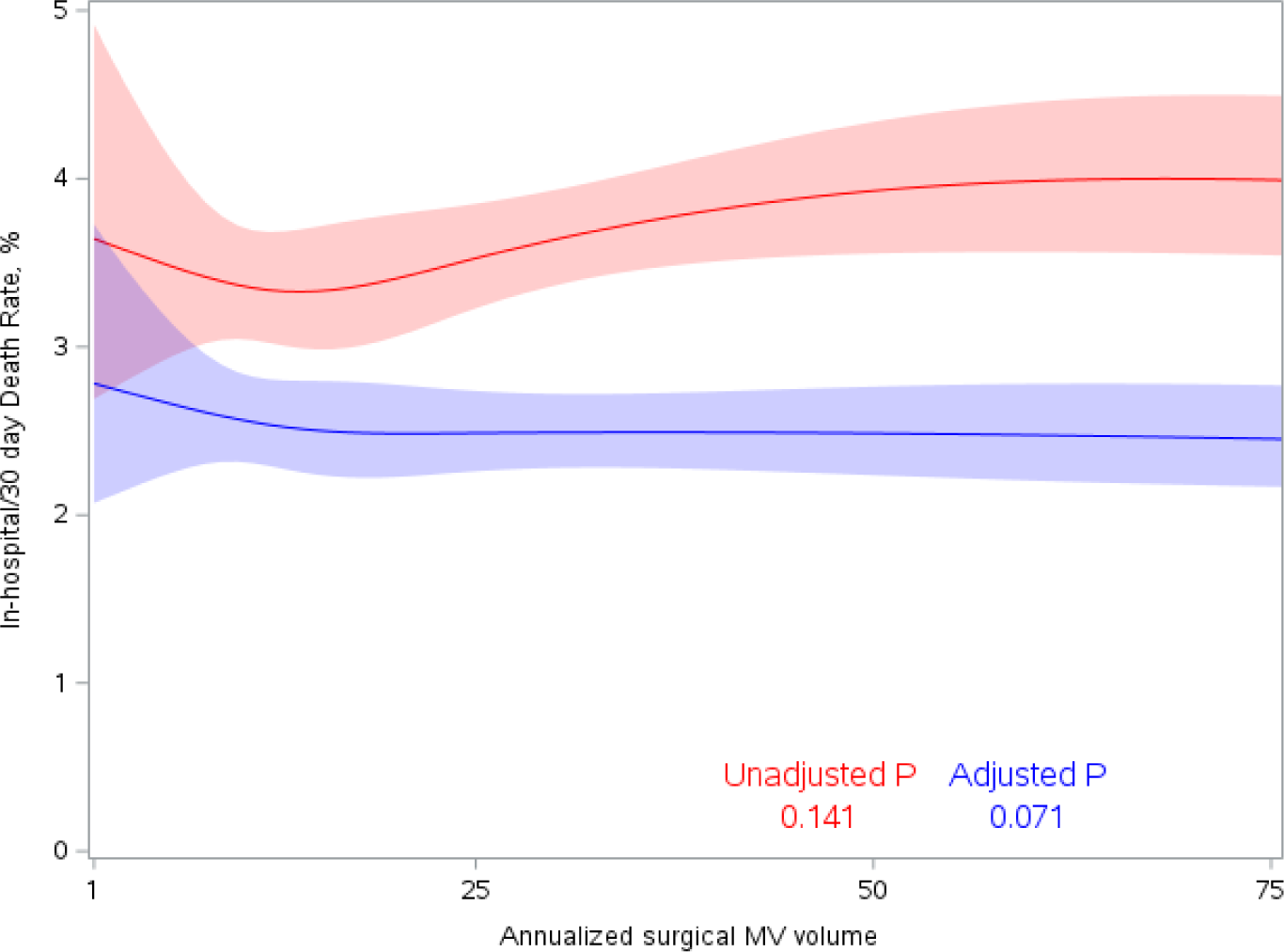
In-hospital or 30 day TEER death by annualized surgical MV volume.

**Table 2.**
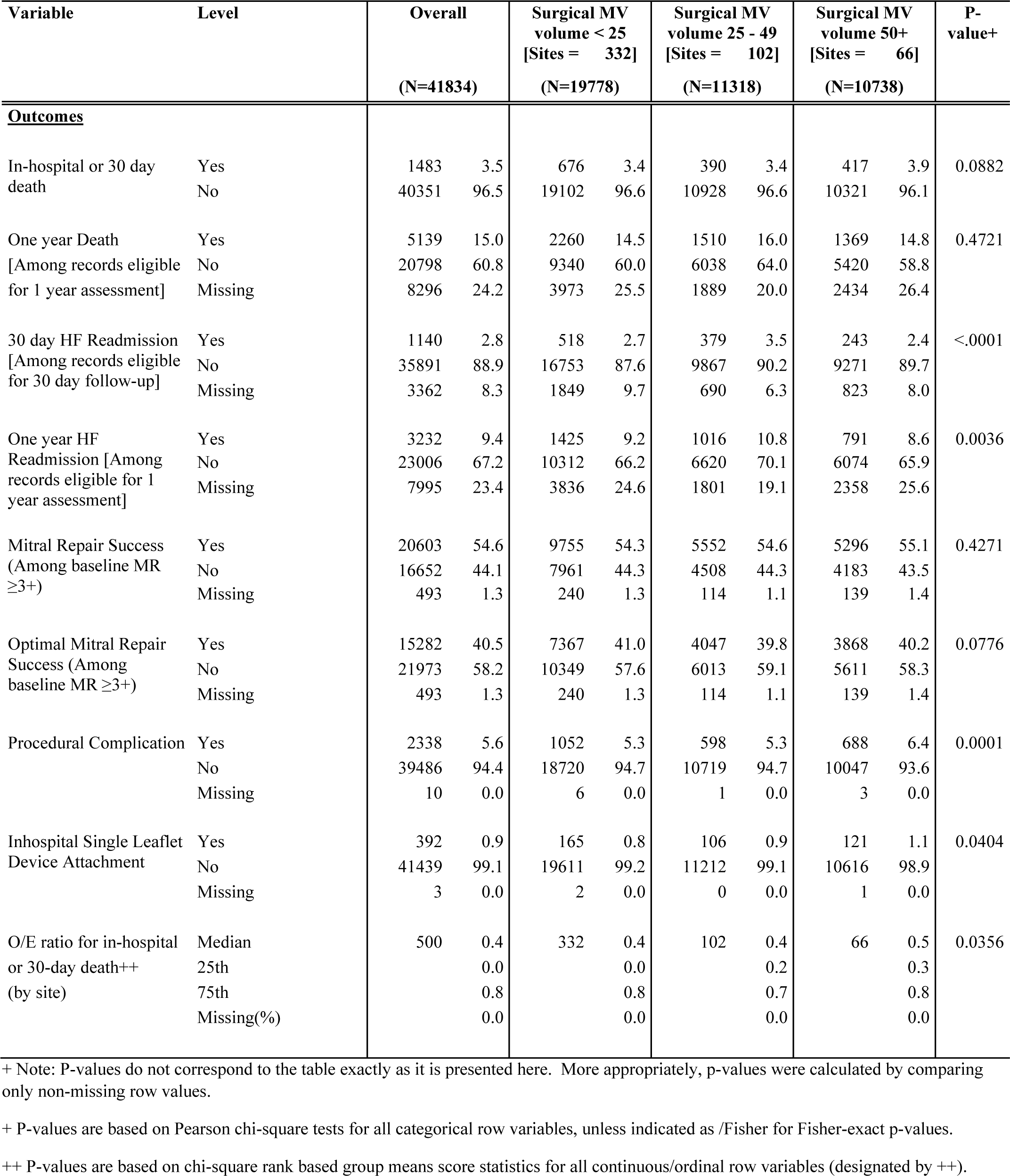
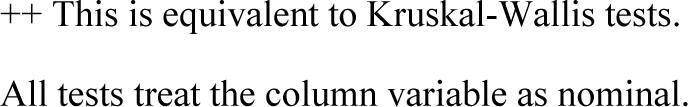
Outcomes by annualized surgical MV volume (raw data).

Figure 2B shows restricted cubic spline analysis for one-year mortality with no statistically significant difference on unadjusted (p=0.118) analysis; however, when adjusted for clinical and demographic variables there was a statistically significant difference (p=0.027) with lower mortality for higher surgical volume centers. One-year mortality was 15.0% overall and also was not significantly different across MV surgical volumes at 14.5%, 16.0% and 14.8% (p=0.4721). When adjusted for clinical and demographic variables, the relationship between annualized surgical MV volume and one-year mortality remained non-significant (p=0.333, Supplemental Table 2B).

**Figure 2B.**
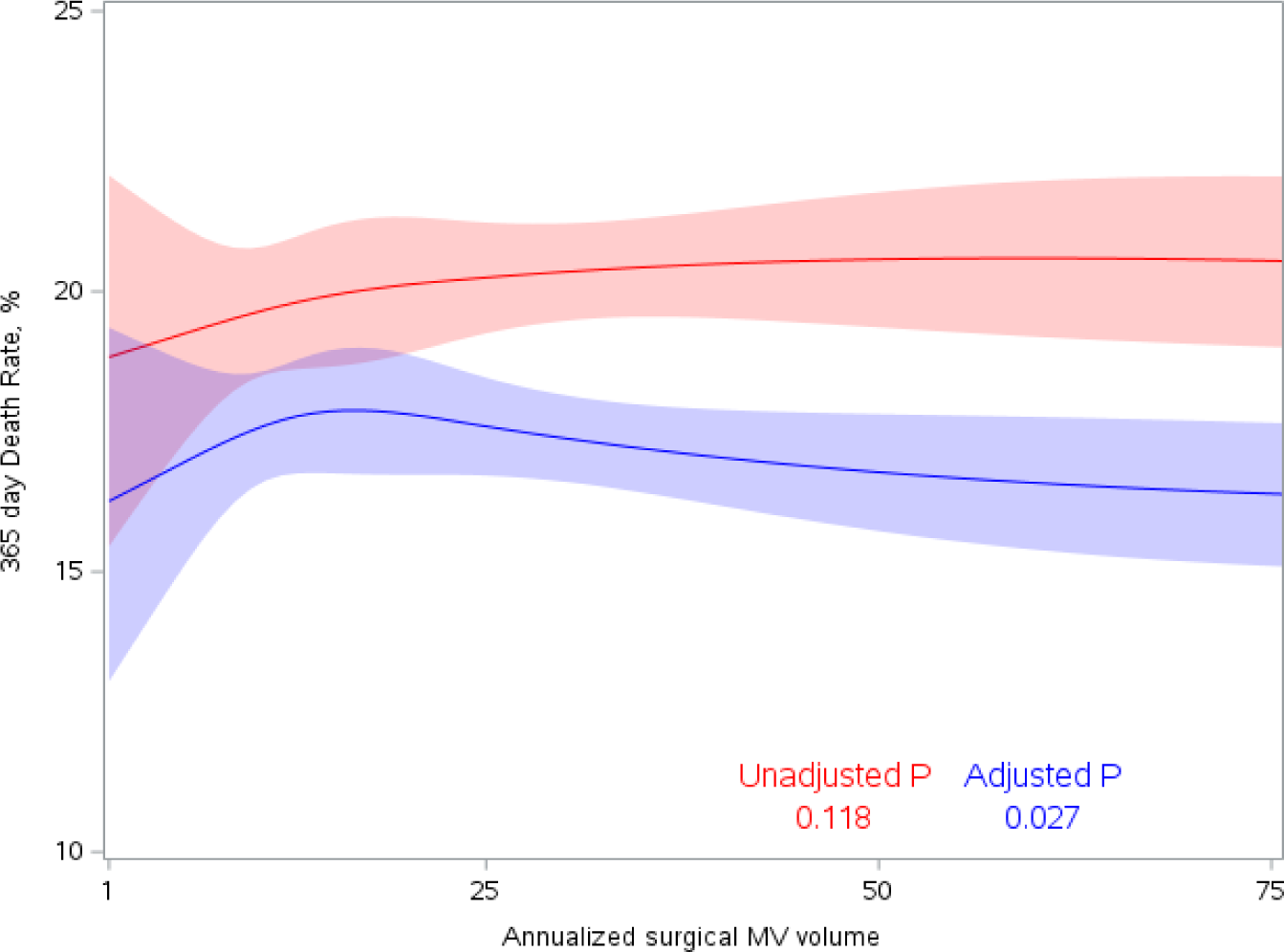
One-year TEER death rate by annualized surgical MV volume.

### HF Readmission (Secondary Endpoint)

Figure 3A shows restricted cubic spline analysis for in-hospital or 30-day HF readmission with no statistically significant difference on unadjusted (p=0.456) or adjusted (p-0.161) analysis. HF readmission at 30-days was 2.8% overall and was statistically significantly different across MV surgical volumes at 2.7%, 3.5% and 2.4% in MV surgical sites with <25, 25-49 and ≥ 50 annualized cases (p<0.001). When adjusted for clinical and demographic variables, the 30-day HF readmission remained statistically significant (p=0.002, Supplemental Table 3A).

**Figure 3A.**
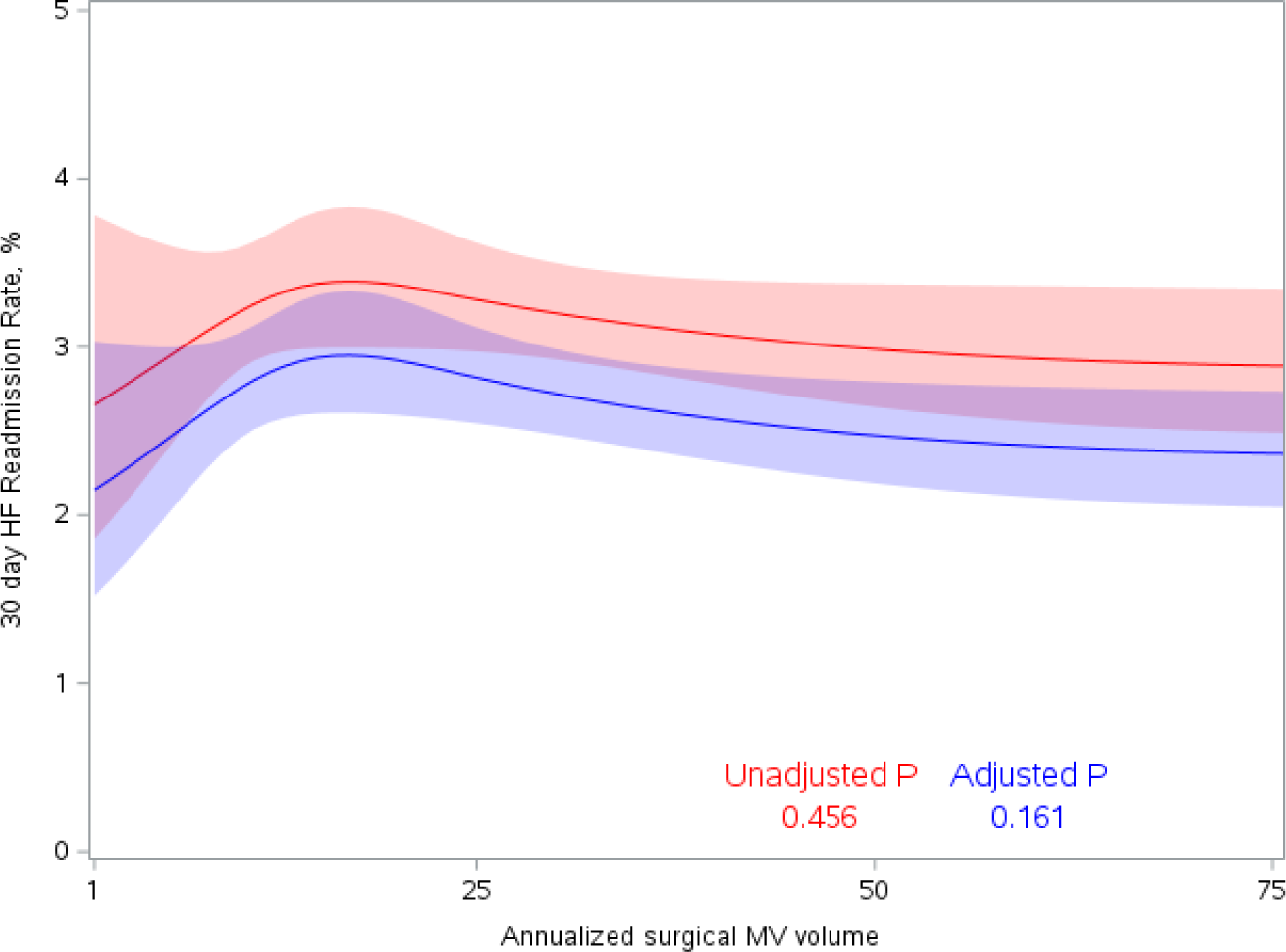
30-day TEER HF readmission by annualized surgical MV volume.

Figure 3B shows restricted cubic spline analysis for one year HF readmission with statistically significant differences on both unadjusted (p=0.017) or adjusted (p-0.015) analysis. HF readmission at one year was 9.4% overall and was statistically significantly different across MV surgical volumes at 9.2%, 10.8% and 8.6% in MV surgical sites with <25, 25-49 and ≥ 50 annualized cases (p=0.0036). When adjusted for clinical and demographic variables, the one-year HF readmission was no longer statistically significant (p=0.149, Supplemental Table 3B).

**Figure 3B.**
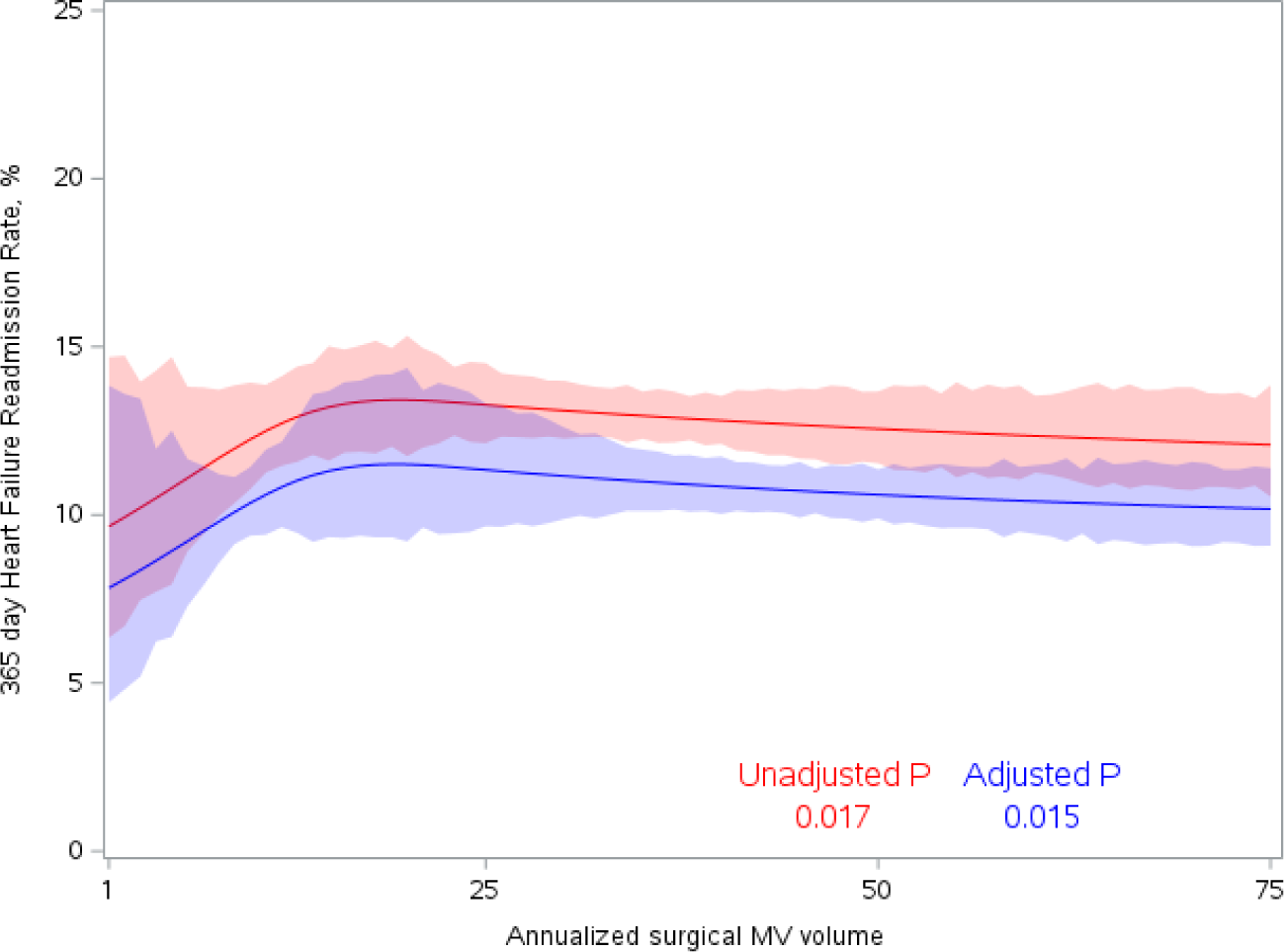
One-year TEER HF readmission by annualized surgical MV volume.

### MR Repair Success (Secondary Endpoint)

Figure 4A shows restricted cubic spline analysis for MR repair success, with no statistically significant differences on unadjusted (p=0.347) or adjusted (p=0.246) analysis. As shown in Table 2, overall MR repair success was 54.6% and was not statistically significantly different across MV surgical site volumes at 54.3%, 54.6% and 55.1% (p=0.4271). These differences remained non-significant on adjusted analysis (p=0.958, Supplement Table 4A). Optimal MR success was 40.5% overall and was not statistically significantly different across sites at 41.0%, 39.8%, 40.2% (p=0.0776). The individual components of MR success included MR grade and mean transmitral pressure gradient. As shown in Figure 5, 94.1% of patients had severe (3+ or 4+) MR at baseline. After the procedure, MR grade was ≤1+ in 64.6%, 2+ in 26.5% and ≥3=+ in 8.9%. Figure 6 shows categories of mean transmitral gradient for each grade of post-TEER MR. After exclusion of missing data, 22,088 patients (59.3%) had mean gradient 0-5 mmHg, 13,830 (37.1%) had mean gradient 5-10 mmHg and 1,337 (3.6%) had mean gradient ≥ 10 mmmHg. The distribution of values across post-TEER MR grades was statistically significantly different from expected values (χ^2^ 526.5, p<0.0001), with mean gradient ≥ 10 mmHg more commonly seen with severe (≥ 3+) MR post-TEER.

**Figure 4.**
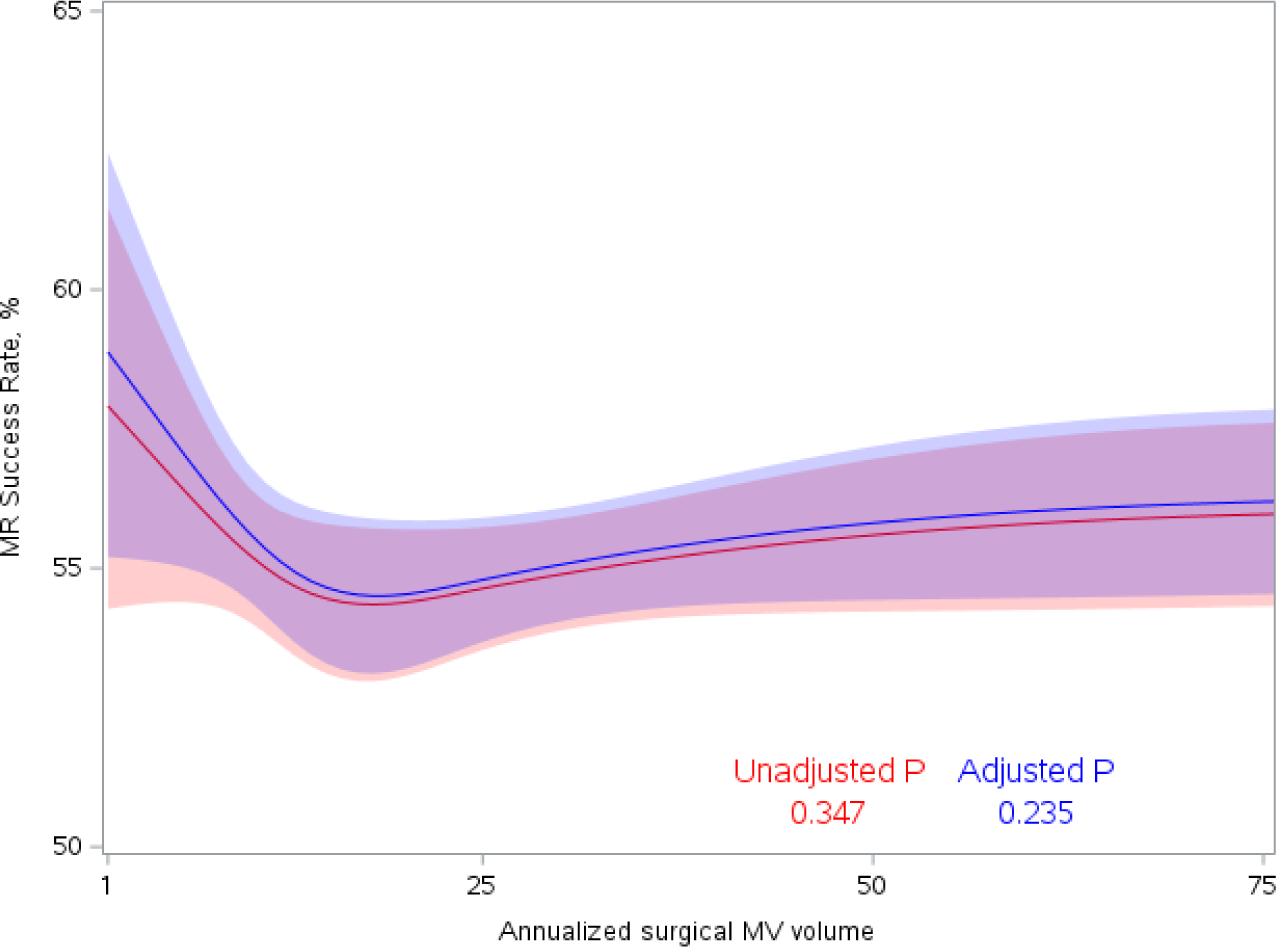
Device success by annualized surgical MV volume.

**Fig. 5.**
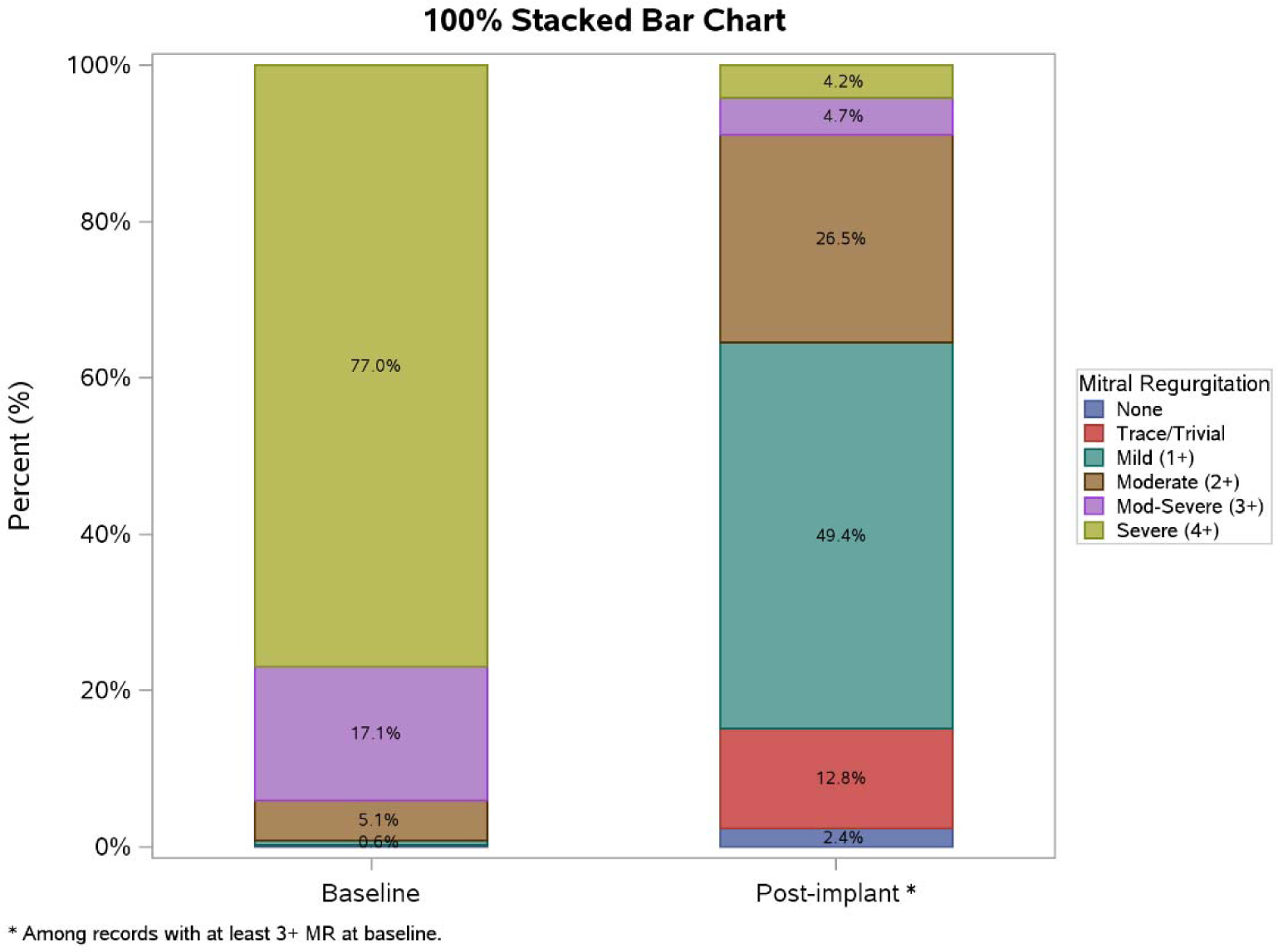
Baseline and post-implant MR grade.

**Fig 6.**
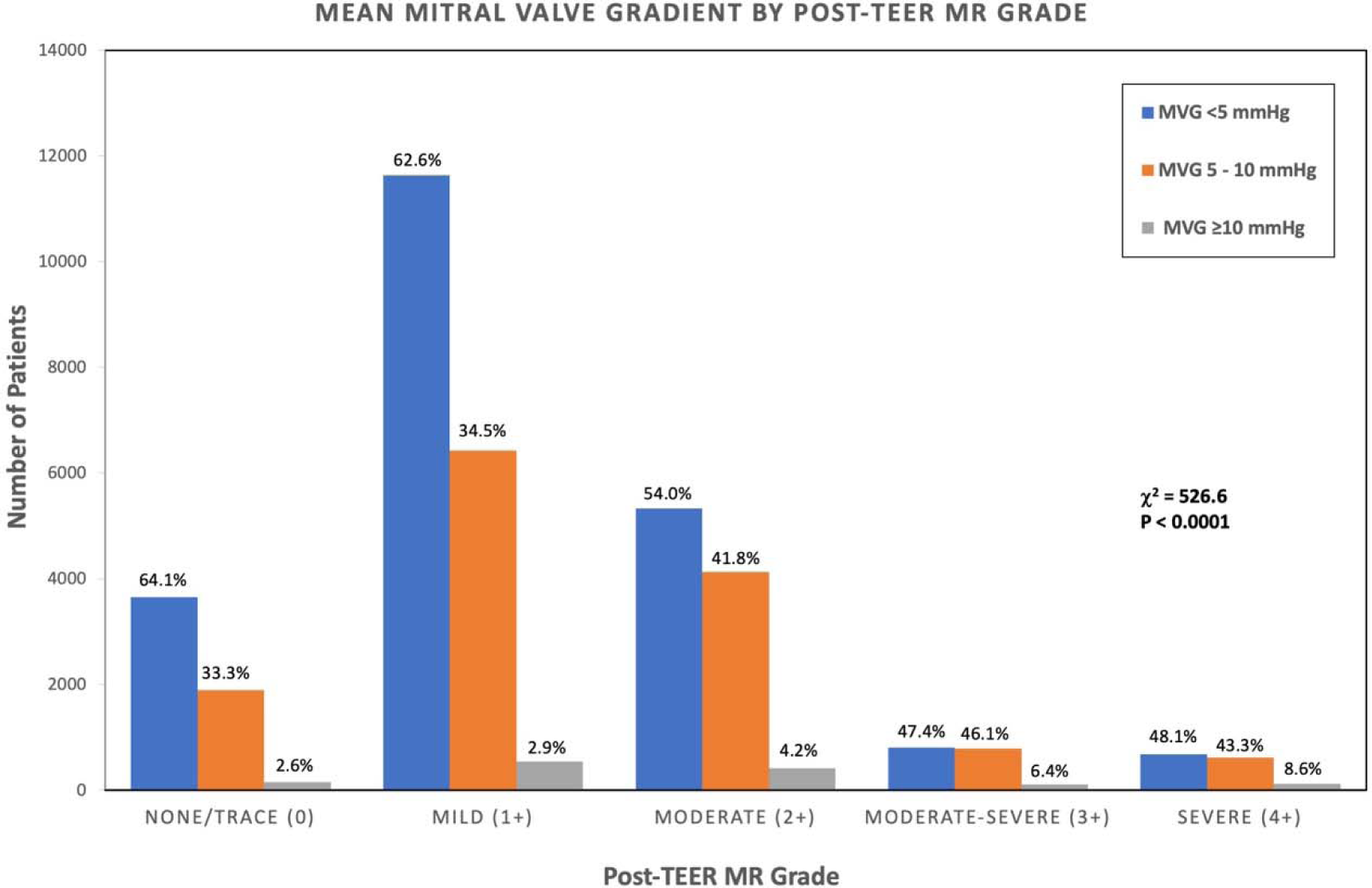
Categories of mitral valve mean gradient by post-TEER MR grade.

### Procedural Complications

Procedural complications were slightly higher in high volume surgical centers (5.3%, 5.3%, 6.4%, p<0.0001). In-hospital single leaflet device attachment was slightly higher in high volume surgical centers (0.8%, 0.9%, 1.1%, p=0.04). Adjustment for demographic and clinical risk factors was not performed.

## Discussion

The study demonstrates several important findings. First, the majority of TEER procedures in the U.S. are done in low volume MV surgery centers (<25 complex MV repairs per year). Only 13% of TEER procedures are performed in high volume MV surgery centers. Second, 30-day TEER outcomes were clinically similar overall for low, intermediate and high volume centers. At 30 days, mortality was 3.5% and was not different across surgical volumes either as a continuous variable by cubic spline, nor when analyzed categorically by pre-defined surgical volume groups. HF readmission at 30 days was 2.8% and was not statistically significantly difference across surgical volumes by cubic spline analysis. Moreover, there was no statistically significant difference in MV repair success by TEER at 30-days. These data suggest that TEER can be performed safely in low surgical volume centers with 30-day outcomes that are similar to those are high surgical volume centers. However, at one-year, there was a statistically significant improvement in mortality by cubic spline analysis when adjusted for clinical and demographic variables (p=0.027). At one-year, HF readmission was statistically significantly lower with higher surgical volumes by cubic spline analysis (unadjusted and adjusted). Thus, the similar outcomes seen at 30-days were not sustained at one-year. Potential explanations include patient selection, unmeasured variables or variables not captured in the database, closer followup by heart team personnel and availability of advanced therapies/consultants for heart failure, kidney failure, and other complications at high volume centers. Of note, the raw mortality of 15.0% at one-year is lower than that reported in earlier series (11,12), suggesting that TEER outcomes may be improving over time. Factors contributing to this cannot be ascertained in this study but could include patient selection, indication creep toward lower risk patients, improvement in technique/device design (13,14), group learning and increasing operator experience.

A somewhat surprising finding of this study was the lower rate of MV repair success (54.6%) or optimal MV repair success (40.5%) compared to previous literature. This is due to use of different definitions. The definition of MV repair success in the ACC/STS TVT national registry includes post-implant MR grade and transmitral mean pressure gradient. This is similar to the Mitral Valve Academic Research Consortium (MVARC), (15) definition of device success (rather than MV repair success). Device success included absence of procedural mortality or stroke, proper device positioning, freedom from unplanned surgical or interventional procedures and “reduction of MR to either optimal or acceptable levels without significant mitral stenosis (i.e., post-procedure MV area is > 1.5 cm^2^ with a transmitral gradient <5 mm Hg), and with no greater than mild (1+) MR.” However, most prior studies have focused only on MR reduction without including mitral gradient. In this series, ≥3+ MR at 30 days was present in 8.9% of patients, similar to other recent reports (5,12-14) and better than that reported in the EVEREST II randomized trial of TEER compared to surgical repair (1). Makkar, et al (16) recently reported the efficacy of TEER in 19,088 patients with primary MR and showed similar findings with an overall success of 52.4%. In that study, an unsuccessful procedure (> moderate MR and mean gradient ≥ 10 mmHg) was associated with a high one-year mortality (26.7%). Conversely, there was a step-wise decrease in mortality with different degrees of MR/mean gradient combinations with an 11.4% one-year mortality in patients with ≤ mild MR and mean gradient ≤ 5 mmHg.

There has been conflicting evidence on the importance of resting gradient post-TEER. In this large ACC/STS TVT national registry, 41% of patients post-TEER had a resting transmitral mean gradient of ≥5 mmHg which is considered to be stenotic (17). In the COAPT trial of TEER in secondary MR, resting gradient had no impact on outcomes in patients with symptomatic HFrEF and severe MR (18). Patzelt et al (19) found an increase in the combined outcomes of endpoint of death, HF hospitalization, mitral surgery, repeat hospitalization, repeat TEER or LV assist device with elevated mitral gradient post-TEER in primary but not secondary MR. Neuss, et al (20) also studied a mixed population of primary and secondary MR and found an increased risk of the same combined endpoint with higher gradients, but did not distinguish between underlying etiology. As noted above Makkar, et al (16), showed clearly that mean gradient in combination with residual MR, is a strong predictor of one-year mortality in patients undergoing TEER for primary MR. In patients at prohibitive surgical risk due to age and multiple comorbidities, an elevated resting gradient may be tolerated as these patients are often sedentary. However, as TEER moves toward a lower risk population eligible for surgery, as in the REPAIR (NCT04198870) or CTSN Primary trials (NCT05051033), resting transmitral gradients could increase with activity and result in functional limitations. The importance of exercise-induced increase in mitral gradient and pulmonary artery systolic pressure has been emphasized the assessment of mitral stenosis (17). However, only one small study has assessed the effects of exercise on mean mitral gradient after TEER (21). The ACC/STS TVT national registry does not include evaluation of physiological function after TEER, so this remains an important area for future studies.

Prior studies have shown improved TEER outcomes with institutional and operator volume. Chhatriwalla et al (4) evaluated cumulative institutional TEER volume in the ACC/STS TVT national registry in 12334 patients 275 sites from Nov 2013-Sept 2017. Optimal procedural success increased across tertiles of institutional case experience (62.0%, 65.5%, and 72.5%, respectively; p < 0.001) and was statistically significantly better with cumulative case volumes > 50. Chhattriwalla et al (5) also reported operator volumes in 14,923 cases from 290 sites from Nov 2013 to March 2018. Statistically significant improvements in optimal procedural success were seen across pre-specified tertiles (1-25, 26-50 and > 50 cases). Both of these studies included a mixture primary and secondary MR. Given the robustness of these prior studies, our study did not repeat the evaluation of institutional or operator volume or potential interaction with hospital MV surgical volumes. Recently, Lowenstern et al (22) reported that the introduction of TEER has not reduced the volume of surgical MV repair but is associated with fewer high risk surgical cases and improved mortality out to 5-years. Badhwar et al (23) in a contemporary analysis of surgical MV repair for primary MR found that the risk of 30-day mortality was <1% in two-thirds of patients and 2.68% in the 95th percentile. In our series, the predicted risk of mortality was 5.0 % overall, which places them well over the 95th percentile for MV surgical repair.

### Limitations

The ACC/STS TVT national registry is a retrospective observational database. Although definitions are pre-specified, there are missing data and unmeasured variables that could influence results. Echocardiographic data are site-reported and were not adjudicated by an expert core laboratory. It is possible that some patients considered to be prohibitive surgical risk at lower MV surgical volume sites might have been referred for surgical MV repair at higher MV surgical volume centers where minimally invasive techniques and additional procedures to address tricuspid regurgitation (24) or atrial fibrillation (25) could be performed. While this possibility is speculative, supplemental Fig 1 shows the ratio of MV surgery to TEER in this study. A ratio < 1 (more TEER procedures than surgical repair) was present in 132 sites (26.4%). This seems to be quite high given that the gold standard for treatment of severe MR due to flail leaflet or prolapse is surgical repair and TEER is approved only for such patients when they are at prohibitive risk of surgery. Current guidelines strongly advocate a multidisciplinary heart team with an experienced MV repair surgeon for clinical decision making in these patients (9,17). Finally, our study evaluated mortality, HF readmission and MV repair success up to one-year followup. Longer term followup to at least 5-years is needed.

## Conclusions

In this large series using the combined ACC/STS TVT national registry and STS database, 30-day outcomes for TEER were similar in terms of mortality, HF readmission and MV repair success across low, intermediate and high volume MV surgical centers. Thus, TEER can be safely performed even in centers with low volumes of complex MV repair. However, at 1-year, mortality and HF readmission improved with higher MV surgical volumes. The reasons for improved outcomes at 1-year cannot be determined from this retrospective registry but are an important target for future investigation.

## Data Availability

This study combined data from both the ACC/STS TVT registry and the STS adult cardiac surgery database which are held at Duke Clinical Research Institute.

## Disclosures

Dr. Grayburn receives research grants/advisory board fees from Abbott Vascular, Boston Scientific, Cardiovalve, Edwards Lifesciences, Medtronic, Neochord, Restore Medical, W.L. Gore and 4C Medical. Dr. Mack has served as a co-principal investigator for Edwards Lifesciences and Abbott trials; and has served as a study chair for Medtronic. Dr Kosinski reported grants from American College of Cardiology. Dr. Szerlip is a speaker and proctor for Edwards LifeSciences, Boston Scientific, and Abbott; is on the advisory board for Boston Scientific; and is on steering committees for Medtronic and Abbott. Dr Smith has received institutional grant support from Edwards Lifesciences, Abbott, and Artivion; has served as a speaker for Edwards Lifesciences, Abbott, Artivion, and Medtronic; and has served on the advisory board for Edwards Lifesciences. Dr Vemulapalli has received grants/contracts from the American College of Cardiology, Society of Thoracic Surgeons, Cytokinetics, Abbott Vascular, National Institutes of Health (R01 and SBIR), and Boston Scientific; and has received consulting fees/been on advisory board with Janssen, American College of Physicians, HeartFlow, and Edwards LifeSciences.

## Acknowledgments

This work was funded by the Baylor Scott & White Foundation and Baylor Scott & White Research Institute. The views expressed in this manuscript represent those of the authors, and do not necessarily represent the official views of either The Society of Thoracic Surgeons or the American College of Cardiology Foundation’s National Cardiovascular Data Registry (NCDR). Learn more about the STS/ACC TVT Registry at www.tvtregistry.org.

---

**Supplemental Fig. 1.**
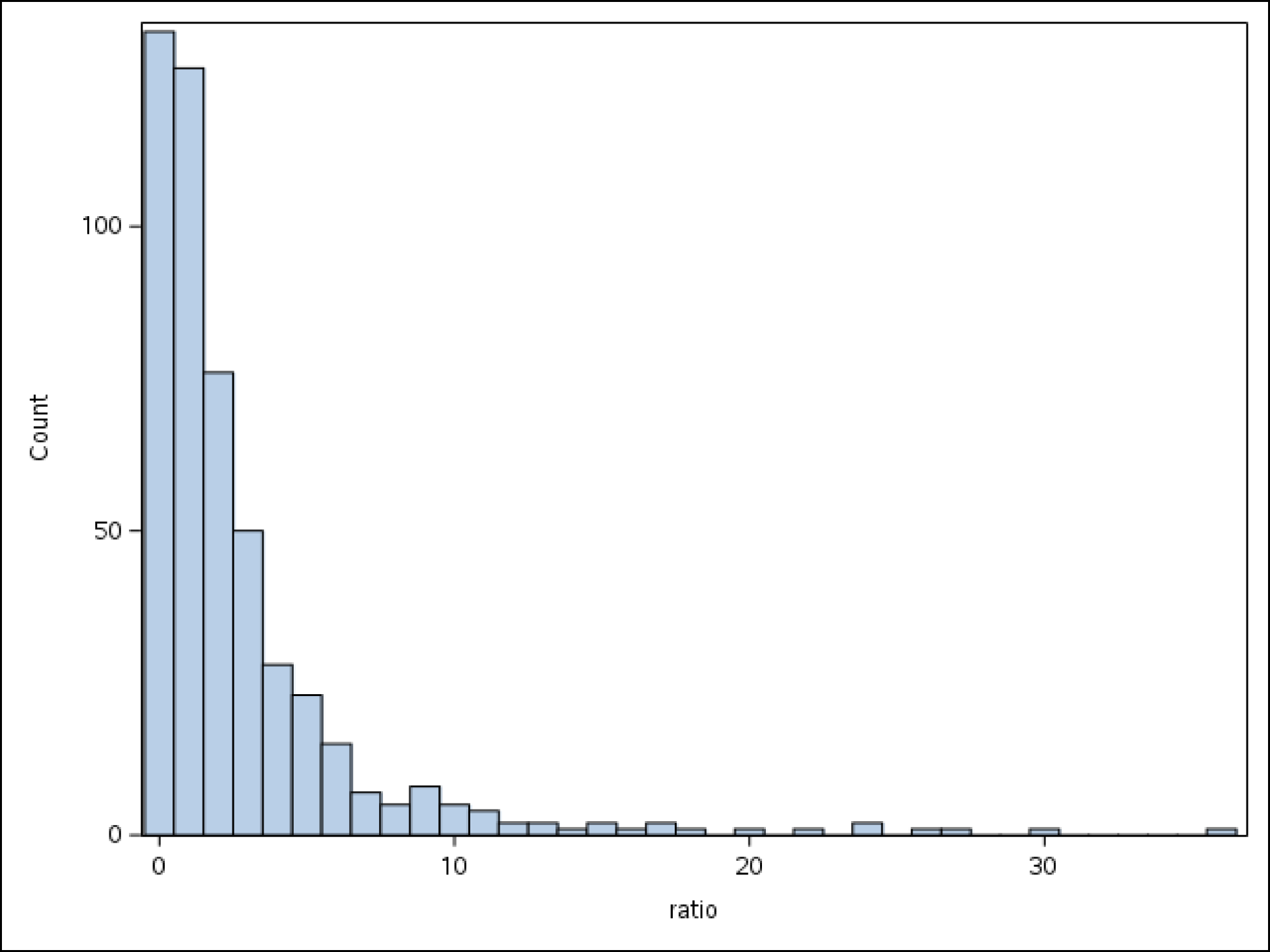
Ratio of surgical MR and TMVr.

Ratio defined as total surgical complex mitral volume divided by total transcatheter mitral repair volume. Ratio > 1 refers to greater surgical volume compared to transcatheter mitral repair volume for a site.

132 sites had surgical mitral volume to transcatheter mitral repair volume ratio of <1.

**Supplemental Table 1.**
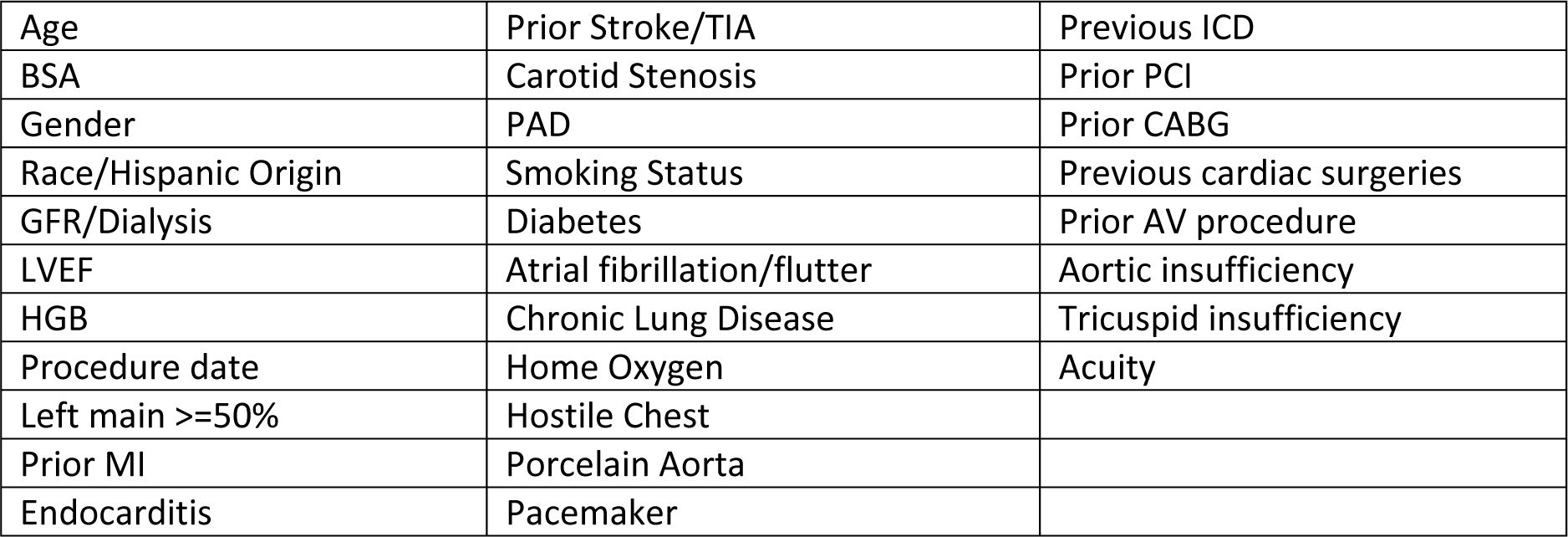
List of variables used for adjustment of mortality and HF readmission endpoints.

|  |  |  |
| --- | --- | --- |
| Age | Prior Stroke/TIA | Previous ICD |
| BSA | Carotid Stenosis | Prior PCI |
| Gender | PAD | Prior CABG |
| Race/Hispanic Origin | Smoking Status | Previous cardiac surgeries |
| GFR/Dialysis | Diabetes | Prior AV procedure |
| LVEF | Atrial fibrillation/flutter | Aortic insufficiency |
| HGB | Chronic Lung Disease | Tricuspid insufficiency |
| Procedure date | Home Oxygen | Acuity |
| Left main >=50% | Hostile Chest |  |
| Prior MI | Porcelain Aorta |  |
| Endocarditis | Pacemaker |  |

**Supplemental Table 2A.** Adjusted association between annualized surgical MV volume and in-hospital/30 day death.

|  | *Adjusted Rate (95% CI)* | *Adjusted OR (95% CI)* | *Adjusted p-value* |
| --- | --- | --- | --- |
| < 25 | 2.57 (2.33, 2.83) | 1.05 (0.90, 1.22) | 0.552 |
| 25 - 49 | 2.37 (2.09, 2.70) | 0.96 (0.81, 1.14) |  |
| 50+ | 2.46 (2.16, 2.81) | Reference |  |

**Supplemental Table 2B.**
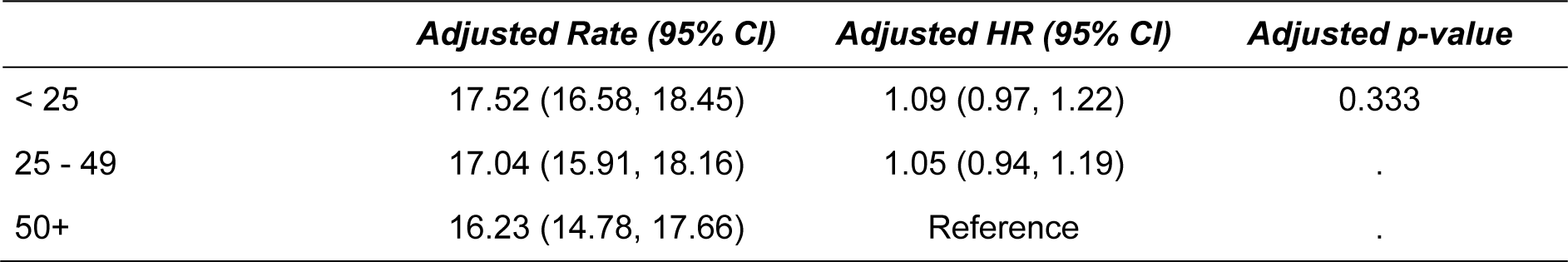
Adjusted association between annualized surgical MV volume and one-year death.

**Supplemental Table 3A.** Adjusted association between annualized surgical MV volume and 30 day HF Readmission.

|  | *Adjusted Rate (95% CI)* | *Adjusted OR (95% CI)* | *Adjusted p-value* |
| --- | --- | --- | --- |
| < 25 | 2.62 (2.36, 2.91) | 1.22 (1.01, 1.48) | 0.002 |
| 25 - 49 | 3.10 (2.72, 3.53) | 1.45 (1.18, 1.78) | . |
| 50+ | 2.16 (1.83, 2.54) | Reference | . |

**Supplemental Table 3B.**
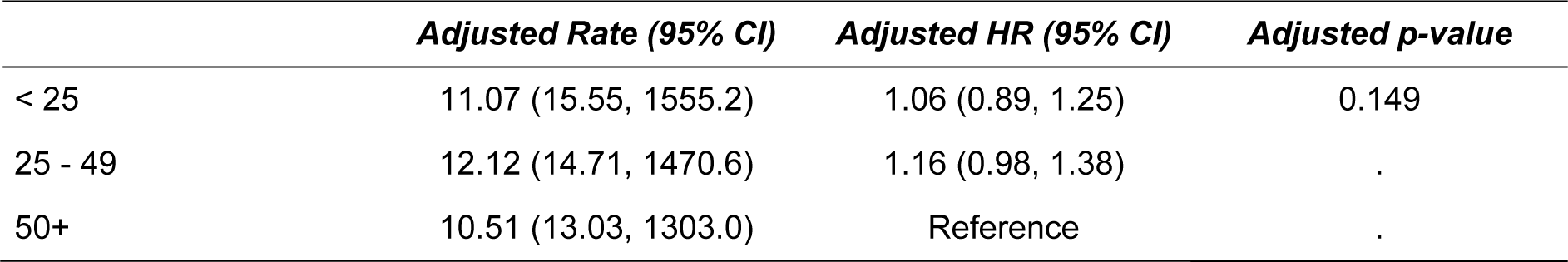
Adjusted association between annualized surgical MV volume and one-year HF readmission.

**Supplemental Table 4.** Adjusted association between annualized surgical MV volume and TEER repair success.

|  | *Adjusted Rate (95% CI)* | *Adjusted OR (95% CI)* | *Adjusted p-value* |
| --- | --- | --- | --- |
| < 25 | 55.46 (54.34, 56.58) | 0.99 (0.91, 1.08) | 0.958 |
| 25 - 49 | 55.75 (54.10, 57.39) | 1.00 (0.91, 1.11) | . |
| 50+ | 55.64 (53.79, 57.48) | Reference | . |

## Notes

### Clinical Trial

This is not a prospective clinical trial. It is a retrospective analysis with a pre-specified statistical analysis plan using data from the TVT/STS NCDR Registry and the STS Adult Cardiac Surgery Database.

### Author Declarations

The STS/TVT Registry has approval through the Advarra IRB and this analysis was granted a waiver by the Duke University Institutional Review Board.

